# Physiotherapists’ use of aerobic exercise during stroke rehabilitation: a qualitative study using chart-stimulated recall

**DOI:** 10.1101/2023.12.13.23299927

**Authors:** Azadeh Barzideh, Augustine Joshua Devasahayam, Ada Tang, Elizabeth Inness, Susan Marzolini, Sarah Munce, Kathryn M Sibley, Avril Mansfield

**Affiliations:** Rehabilitation Sciences Institute, University of Toronto, Toronto, Canada; KITE-Toronto Rehabilitation Institute, University Health Network, Toronto, Canada; Department of Physical Therapy, University of Toronto, Toronto, Canada; Evaluative Clinical Sciences, Hurvitz Brain Sciences Program, Sunnybrook Research Institute, Toronto, Canada; Department of Occupational Science and Occupational Therapy, University of Toronto, Toronto, Canada; Institute of Health Policy, Management and Evaluation, University of Toronto, Toronto, Canada; School of Rehabilitation Science, Faculty of Health Sciences, McMaster University, Hamilton, Canada; Community Health Sciences, University of Manitoba, Winnipeg, Canada

**Author notes:** **Corresponding author:** Azadeh Barzideh; address: 550 University Ave, Toronto, ON, M5G 2A2. **Funding:** This study was supported by grants from the Canadian Institutes of Health Research (CIHR PJT-148906 & PJT-173472). **Declaration of interest statement:** The authors report there are no competing interests to declare.

**Keywords:** Stroke, Rehabilitation, Aerobic exercise, Thematic analysis, Physiotherapists

## Abstract

**Purpose:** We aimed to explore the factors that affect physiotherapists’ use of AE during stroke rehabilitation for people with stroke.

**Material and methods:** We conducted a qualitative descriptive study using thematic analysis informed by a pragmatic worldview. Physiotherapists attended one on one semi-structured interviews to answer some general questions about aerobic exercise and then discussed the charts of their 4 most recently discharged patients. Both deductive and inductive coding were used for analysis.

**Results:** Ten physiotherapists participated. Healthcare policies and limited resources were mostly discussed in general questions while specific profiles of patients with stroke, their goals and preferences were mostly discussed in patient specific questions. Three themes were identified:1) physiotherapists’ perspectives and practices around aerobic exercise; 2) profiles of people with stroke, as well as their goals and their exercise modality preferences; and 3) influence of health system priorities, rehabilitation intensity policy, and resources.

**Conclusions:** Physiotherapists’ behaviours regarding use of aerobic exercise for people with stroke are not a binary behaviour of prescribing or not prescribing aerobic exercise. Their behaviours are better understood on a continuum; between two ends of not prescribing aerobic exercise, and prescribing aerobic exercise with defined intensity, duration, and frequency.

## INTRODUCTION

People with stroke often have low cardiorespiratory fitness, which makes it challenging for them to complete daily activities, such as carrying groceries and doing housework [1]. Evidence shows that aerobic exercise post-stroke improves cardiorespiratory fitness, and cardiovascular risk factors by decreasing systolic blood pressure, fasting glucose, fasting insulin levels, and increasing high-density lipoprotein cholesterol levels [2]. Furthermore, aerobic exercise can improve cognition [3], walking capacity [4] and facilitate the completion of activities of daily living[5]. Therefore, clinical guidelines recommend the prescription of aerobic exercise during routine stroke rehabilitation[6].

Seventy eight percent of public inpatient and outpatient stroke rehabilitation programs surveyed across Canada report including an aerobic exercise component [7]. Additionally, 77% of physiotherapists working with stroke patients report prescribing aerobic exercise [8]. However, the number of patients who participate in aerobic exercise during stroke rehabilitation is low. For example, in one Canadian inpatient rehabilitation setting where the treating physiotherapists were responsible for performing submaximal exercise testing and prescribing aerobic exercise to people with stroke, only 37-42% of stroke patients participated in exercise [9].

Numerous studies have examined what prevents and facilitates physiotherapists implementing aerobic exercise during rehabilitation [7,8,10,11]. In a recent scoping review, the barriers and facilitators to aerobic exercise implementation in stroke rehabilitation from physiotherapists’ perspectives were investigated [12]. This review reported barriers related to “environmental context and resources” (e.g., lack of equipment, time, staff), insufficient “knowledge” and “skills” (e.g., safe aerobic exercise prescription and implementation), “beliefs about capabilities” (e.g., uncertainty about exercise intensity and screening tools), and “professional role and identity” (e.g., aerobic exercise is not a priority for physiotherapists). Alternatively, having access to and continued education in structured aerobic exercise programs and safety monitoring were identified as facilitators to aerobic exercise implementation [12]. Another recent systematic review [13] found that professionals’ self-efficacy and knowledge about stroke and patients’ needs are among the factors influencing implementation of aerobic exercise for people with stroke [13]. Additionally, communication and collaboration within and between organisations to aid knowledge sharing between professions and services and resources such as equipment, staff and training are other key factors that influence aerobic exercise implementation after stroke [13]. Concerns for patients’ safety in both assessing the aerobic capacity (using submaximal exercise testing) and performing aerobic exercise are repeatedly reported in studies [8,10,14].

Studies exploring physiotherapists’ and stroke program managers’ perspectives on barriers and facilitators to including aerobic exercise in stroke rehabilitation programming have involved mostly the collection of data via surveys, one-on-one semi structured interviews and focus groups [7,8,10,14,15]. Physiotherapists respond to the survey and interview questions by giving their general perceptions about their aerobic exercise prescription practices. There may be discrepancies between clinicians’ practice and their general perceptions of their practice [16]. Chart stimulated recall (CSR) is a clinical research tool that can provide insights into healthcare professionals’ reasoning for clinical decision making for specific patients, the social and clinical influences on decisions, and discrepancies between clinicians’ real and perceived practice [16]. CSR involves the clinician referring to specific patient charts during interviews to provide information on what affected their decision-making process. In this study, we aimed to explore physiotherapists’ perspectives about using aerobic exercise for individual patients via CSR, to obtain a better understanding of physiotherapists’ clinical reasoning and social and clinical factors affecting their decisions for each specific case.

## METHOD

### Study design & theoretical framework

We conducted a theoretically informed qualitative descriptive study [17] guided by a pragmatic worldview [18]. Two theoretical frameworks informed this study, namely, the Capability Opportunity Motivation Behaviour (COM-B; [19]) model and Theoretical Domain Framework (TDF; [20]). COM-B provides a model for understanding behaviours with a goal of behaviour change. TDF integrates 33 theories and 128 key theoretical constructs related to behaviour change and synthesises them into a single framework to assess implementation and other behavioural problems and inform intervention design. TDF consists of 14 different domains [21]. The standards for reporting qualitative research (SRQR) [22] were followed in reporting this study.

### Participant selection and setting

We invited all physiotherapists working with people with stroke in four urban rehabilitation hospitals in Ontario, Canada (Toronto Rehabilitation Institute – University Centre, Toronto Rehabilitation Institute – Rumsey Centre, St. John’s Rehabilitation – Sunnybrook Health Sciences Centre, and Hamilton Health Sciences – Regional Rehabilitation Program) to participate in the study. These sites were chosen because they all offered structured aerobic programs through our previous research study [23] and were provided with materials and resources to run aerobic exercise groups post-stroke. Physiotherapists in these centres received training in submaximal aerobic capacity testing and exercise prescription for individuals with stroke [23]. Therefore, it is expected that physiotherapists at these centres will not experience the commonly-reported barriers of time, knowledge, and resources to implementing new healthcare practices. All sites provide both in-patient and out-patient stroke rehabilitation, except for the Rumsey Centre site, which only offers out-patient rehabilitation.

The physiotherapist participants were identified by clinical managers and their colleagues at each site. The principal investigator or site lead then contacted the physiotherapists via e-mail and invited them to contact the research assistant if they were interested in participating. The research assistant obtained written informed consent. The research ethics boards of all participating sites approved the study (University Health Network protocol number: 20-5695, Sunnybrook Research Institute protocol number: 3605, and Hamilton Health Sciences protocol number: 11523).

### Data collection

Qualitative data collection with participants occurred via one-on-one semi-structured interviews using CSR. The interview guide is provided in the supplementary Appendix 1. These interviews were conducted by the PhD candidate (AB) via Microsoft Teams (Version 1.5.00.17656, Redmond, Washington, USA) during which participants were asked general questions about aerobic exercise and then to discuss charts from their four most recently discharged patients. Physiotherapists were asked to have the chart available to refer to during the interview. All the charts were discussed during the interview regardless of whether the patient was prescribed aerobic exercise or not. The patients were not interviewed. Another research assistant was also present during the interviews and took notes on participants’ expressions, body language and key data from the interview from their own perspective. The interviews began with providing a definition of aerobic exercise to the physiotherapists. Based on the American College of Sports Medicine’s definition, we explained that aerobic exercise is planned, structured, and repetitive bodily movement done to improve and/or maintain cardiorespiratory fitness with certain threshold criteria (e.g., specific frequency, duration, and intensity) [24,25]. General questions, guided by COM-B, about the physiotherapist’s capability, opportunity, motivation, and general behaviours around aerobic exercise use post-stroke. Following this, the next set of questions related to decision making for specific patients via CSR, with questions and prompts on patient capability and motivation, and physiotherapist capability and opportunity for this specific patient. The interview guide was pilot tested with two physiotherapists from an acquired brain injury unit who were not eligible to participate in the study (but had experience prescribing aerobic exercise) for feedback regarding clarity of the questions; modifications were made as indicated. All interviews were audio-recorded and transcribed verbatim. The interviewer (AB) wrote field notes after each interview. Each interview lasted approximately 1 hour.

To determine our sample size, we relied on a mix of interpretive and pragmatic judgements [26]. For the interpretive judgement, we used information power as the guide for our sample size, which indicates that fewer participants are required when there is more relevant information in the sample [27]. Information power is determined based on broadness of the aim of the study, specificity of the participants’ experience, whether there is an established theory used in the study, quality of dialogue with participants, and the analysis strategy [27].

We compared the criteria for information power to a recent study with similar methodology and aim [28]. This study aimed to explore the perceived barriers and facilitators that influence Australian physiotherapy practices when prescribing strength training with people with stroke during gait rehabilitation and included 1:1 semi structured interviews with physiotherapists. This study reached thematic saturation at 16 interviews. Although the factors affecting information power in our study were similar to this study, we believe our sample held more information. The assumed higher information power in our study is due to the use of CSR and focusing the discussion on 4 specific patient cases with the physiotherapists, while the study on strength training only asked general questions about the perceived barriers and facilitators physiotherapists encounter in practice. Therefore, we concluded that fewer than 16 interviews would be required in our study.

Regarding pragmatic considerations, all 29 physiotherapists at the 4 rehabilitation centres were invited to participate. However, after multiple invitations 17 did not respond, leaving us with interviews from one third of the eligible population. Taking these factors into consideration, we believe we had sufficient number of participants to be able to contribute to the knowledge in the field.

### Data analysis

We used a codebook thematic analysis in this study [29]. A research assistant first transcribed each audio recording verbatim. Researcher AB, who conducted the interviews, listened to the audio records and edited the transcripts to make sure the transcription process was accurate. Data analysis was performed concurrently with data collection. The Framework Method Procedure [30], which allows for a combined deductive-inductive analysis approach was used in this study. Following stages of the Framework Method Procedure, after transcription of data, two members of the team (AB and AM) first read and familiarized themselves with the data. To facilitate the organization and analysis of the qualitative data, the transcripts were then entered into Nvivo (V.12, QSR International, Burlington, Massachusetts, USA), which was also used to facilitate coding the transcripts. Two coders (AB and AM) with different backgrounds (physiotherapy and kinesiology) coded the transcripts separately. After coding every 2-3 transcripts, the coders met to compare the labels applied and agree on a set of codes to apply to all subsequent transcripts (a codebook). The codebook consisted of a table of the parent codes(i.e., the primary TDF domain), sub-codes, definitions of codes, and the example quotes. The final codebook, which was developed after coding the final transcript, was applied to all transcripts. TDF guided the deductive part of data analysis. Concepts that were seen in transcripts that could not be coded deductively were coded inductively. The notes taken by the research assistant and the interviewer were consulted if there was vagueness in the responses to the interview questions.

When all transcripts were coded using the final codebook, AB summarized all the codes into a spreadsheet (i.e., codes and subcodes in rows and participants in columns) for each physiotherapist. AM checked the summarized data in the excel sheet to ensure that all the details from the codes were included. AB and AM took notes on the relationships between the codes when summarizing them making sure the connections and overlaps were not lost. The coders subsequently created themes inductively and refined them several times in consultation with another co-investigator (SM) and created a model (figure 1) to explain the data.

**Figure 1.**
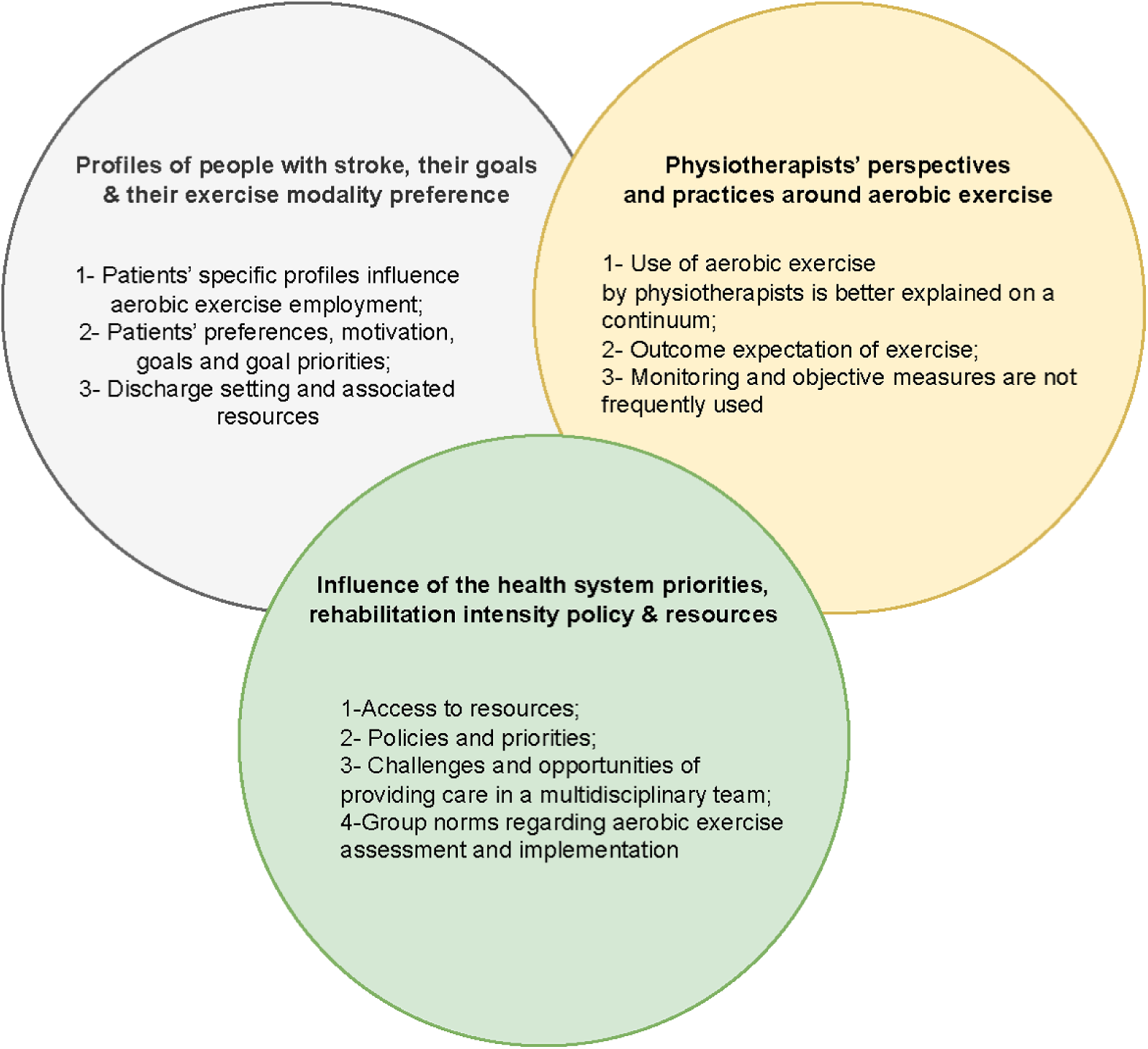
Themes and subthemes contributing to the use of aerobic exercise in stroke rehabilitation.

### Research team

AB, who performed the interviews and was one of the coders, is a female PhD candidate with a background in physiotherapy and completed post-secondary training in Iran, Belgium, and Canada. AM, who was the second coder, is a female Scientist with a background in kinesiology and >12 years of experience as an independent researcher in the field of neurorehabilitation; she is the principal investigator in this study. Both AB and AM performed data analysis and sent the results to SM (Sarah Munce). SM is a female health services and implementation science researcher with approximately 14 years of experience conducting qualitative research; she is a co-investigator on this study. SM checked the results and themes that AB and AM developed and provided feedback.

### Positionality statements

AB acknowledges her standpoint as an internationally educated physiotherapist. While she has not yet practiced in Canada, she has experience working in acute, sub-acute and chronic stroke care hospital units in Iran and Belgium. Therefore, although she is familiar with stroke rehabilitation culture, not practicing in Canada helped her adopt an etic ontological position in this study. It is likely that her physiotherapy background influenced her interview portion as she had similar experiences with many of the physiotherapists in this study when working with people with stroke. Consequently, there were times she did not probe the physiotherapist with more questions because she believed she understood what the physiotherapist was trying to communicate.

As a scientist with a background in exercise science, and research expertise in cardiorespiratory exercise post-stroke, AM has significant knowledge of exercise prescription post-stroke. She has conducted several studies of exercise post-stroke in collaboration with physiotherapists at one site, and previously led the PROPEL trial at all sites. This gives her something of an ‘insider’ perspective in terms of how people with stroke are prescribed exercise; this insider perspective gave her additional context around the setting and physiotherapists’ workflow that influenced data interpretation. However, her position as a scientist means she is disconnected from the clinical teams and, therefore, her primary position is that of an ‘outsider.

### Rigor of the qualitative inquiry

Rigor or data trustworthiness in qualitative research comprises of both validity and reliability of the data [31]. In order to achieve rigor in this study the following methods were used:

- Development of a coding system: as described earlier, a codebook was developed to ensure that the codes and their meanings were the same between the coders. This enhanced both the validity and the reliability of our findings [31].
- Reflexivity or self-reflection: AB kept track of her experiences, reactions, and any potential impact on each interview in a word document for each interviewee [32].
- Triangulation: entails the use of different methods, investigators, data sources, and theories in order to answer the same question and obtain corroborating evidence [31,33]. Our participants were recruited from different hospitals across Ontario providing variation in the sources of data for triangulation. Additionally, triangulation was achieved at the investigator level as AB and AM went through and analysed the data separately before merging the findings and developing themes. AB developed the themes and AM checked the themes and the resulting models (figure 1) explaining the data to make sure no themes were missing. SM later on checked the codes and theme labels to agree on the final theme labels.

Credibility was achieved by the way of development of a codebook, reflexivity and triangulation [31]. Data transferability or generalizability was reached by providing a rich description of the study sample [34]. Confirmability was attained using triangulation and leaving an audit trail between the raw data and interpretations [31,34]. Finally, dependability of the data was obtained by investigator triangulation; meaning three investigators independently reviewed the data and discussed the resulting themes if discrepancies or missing information were detected [34].

## RESULTS

Twenty-nine physiotherapists from 4 hospitals were invited to participate in the study. Twelve physiotherapists initially agreed to participate and signed the consent forms, and the remaining 17 physiotherapists did not respond to the invitations. Two physiotherapists who consented to participate did not complete the interview as they were unable to find time to schedule it. Eight female physiotherapists and 2 male physiotherapists attended the interviews, which were conducted from September 2021 to October 2022. The physiotherapists had an average of 14 years of practice as a physiotherapist (min-max 1 to 30 years) and 9 years in stroke rehabilitation specifically (min-max 1 to 21 years). Names used for physiotherapists are pseudonyms chosen randomly and do not match their real names. Table 1 shows the cases of each physiotherapist and the reasons behind their clinical decision making regarding the use/not use of aerobic exercise.

**Table 1.** Cases with stroke discussed with physiotherapists (PT). Reasons for physiotherapist behaviour regarding aerobic exercise use or lack thereof, for each case. Whether aerobic exercise was employed for each patient or not was based on the PTs’ response to the question “was this patient prescribed aerobic exercise as part of their rehabilitation?” during the interview. Goals were defined in the patient-physiotherapist conversation at the beginning of the physiotherapy sessions.

| PT | Patient | Aerobic exercise | Reason |
| --- | --- | --- | --- |
| Jennifer | 1 | N | Not relevant to the patient's goals* |
|  | 2 | N | Not relevant to the patient's goals |
|  | 3 | N | Not relevant to the patient's goals |
|  | 4 | N | Not relevant to the patient's goals |
| Yvan | 1 | N | Patient low tolerance, lack of time. |
|  | 2 | N | Caseload management, lack of time, PT expected that functional exercises would benefit cardiorespiratory fitness. |
|  | 3 | N | Patient's medical condition, PT expected that functional exercises would benefit cardiorespiratory fitness . |
|  | 4 | N | Not relevant to the patient's goals, staff shortage. |
| Syed | 1 | Y | Patient's profile (young, motivated, no cognitive issues, no cardiac complexity) |
|  | 2 | Y | PT expected aerobic exercise to prevent recurrent stroke and improve endurance in spite of unfavorable patient profile (severe cognitive deficit, no motivation, one sided inattention ) |
|  | 3 | N | Did not enter rehab but was evaluated by the PT for home exercise education, Patient's profile (young, no major complication) |
|  | 4 | Y | Patient's profile (no cognitive issues, physically capable to exercise safely) |
| Helen | 1 | Y | Patient's motivation and goals |
|  | 2 | Y | Patient's profile (good tolerance) and goals |
|  | 3 | Y | Patient's rehabilitation needs (strengthening and endurance) |
|  | 4 | Y | Patient's goals |
| Alta | 1 | Y | PT believed everyone should do aerobic exercise. |
|  | 2 | Y | Patient's goals |
|  | 3 | Y | Patient's goals, PT believed everyone should do aerobic exercise. |
|  | 4 | Y | PT believed everyone should do aerobic exercise. |

**Table 1 continued**
| PT | Patient | Aerobic exercise | Reason |
| --- | --- | --- | --- |
| Adele | 1 | Y | Patient's goals and modality preference, Discharge setting and resources available to the patient. |
|  | 2 | Y | Patient's goals and modality preference, Discharge setting and resources available to the patient resources, Following guidelines. |
|  | 3 | Y | Patient's profile (young, functionally high level, sedentary post stroke) and goals, Discharge setting and resources available to the patient. |
|  | 4 | Y | PT expected aerobic exercise to improve endurance , Discharge setting and its resources. |
| Rejean | 1 | N | Patient's cognitive impairment, High risk of falling and non-functional arm |
|  | 2 | N | Patient's medical condition, Lack of resources. |
|  | 3 | N | Patient's fatigue and spasticity. |
|  | 4 | N | Patient's comorbidities. |
| Courtney† | 1 | Y | Patient's profile (physical activity history, young, good functional movements). |
|  | 2 | Y | Patient's profile (physical activity history, young), Discharge setting and its resources. |
|  | 3 | Y | Patient's profile (recent cardiovascular surgery so activity tolerance needed to improve, physical activity history), PT expected aerobic exercise to improve endurance. |
| Marianne | 1 | Y | Patient's profile (physically capable), Rehabilitation needs (deconditioned) and goals. |
|  | 2 | N | Lack of time and goal priority. |
|  | 3 | N | Patient's profile (high functioning), Discharge guidelines. |
|  | 4 | N | Patient's profile (medical conditions) and goals. |
| Brad | 1 | N | Patient's profile (recent cardiovascular surgery, fatigue), Working in a multidisciplinary team. |
|  | 2 | N | Patient's profile (not deconditioned), COVID, Short length of stay. |
|  | 3 | N | Patient's profile (recent cardiovascular surgery, Severely deconditioned), Lack of time, Lack of fitness group, Complicated processes in the hospital. |
|  | 4 | N | Patient's profile (other medical conditions), Lack of time. |

\* This is the case where the patient had the goal to improve endurance, but the physiotherapist did not employ aerobic exercise specifically because they perceived that it was not relevant to the patient's rehabilitation goals.
† This PT worked part-time in their unit and had recently worked with three stroke patients prior to the interview.

### Themes

#### Overview of themes

Overall, three themes related to aerobic exercise use during rehabilitation were identified: 1) physiotherapists’ perspectives and practices around aerobic exercise (consisting of three subthemes of use of aerobic exercise by physiotherapists is better explained on a continuum; outcome expectation of exercise; and monitoring and objective measures are not frequently used); 2) profiles of people with stroke, as well as their goals and their exercise modality preferences (encompassing three subthemes of patients’ specific profiles influence aerobic exercise use; patients’ preferences, motivation, goals and goal priorities; and discharge setting and associated resources); and 3) influence of the health system priorities, rehabilitation intensity policy, and resources (including four subthemes of access to resources; policies and priorities; challenges and opportunities of providing care in a multidisciplinary team; and group norms regarding aerobic exercise assessment and implementation. The influence of the COVID-19 pandemic on aerobic exercise use cut across all themes; these influences are explained separately in more detail after the themes are introduced.

As depicted in Figure 1, factors affecting use of aerobic exercise for people with stroke can be related to the patients, the physiotherapists or the organization, and they overlap with each other. The overlaps show that more than one factor contributed to a behaviour related to the use of aerobic exercise. For example, having a physically/cognitively unsafe patient when a lack of adaptable equipment exists can lead to not employing aerobic exercise.

### Themes and subthemes

*1) Physiotherapists’ perspectives and practices around aerobic exercise*

#### Use of aerobic exercise by physiotherapists is better explained on a continuum

Physiotherapist behaviour could not be simply described as prescribing or not prescribing aerobic exercise for individual patients but was better explained on a continuum. That is, not using aerobic exercise for any clients was on one end of the continuum, and prescribing aerobic exercise for all clients with defined intensity, duration, and frequency to improve cardiorespiratory fitness was on the other end. A range of behaviours was reported in the middle of the continuum. For example, implementing aerobic exercise without a specific prescription, using the aerobic equipment only for warm up or cool down or prescribing low intensity/short duration exercise. Depending on the patient and circumstances, each physiotherapist may exhibit one behaviour for one patient and another behaviour for another patient. Most physiotherapists described intensity of aerobic exercise based on the equipment they used (e.g., level one on a motorized stationary cycle) and not based on patient’s heart rate or rate of perceived exertion as recommended by the aerobic exercise guideline to optimize best practices in care after stroke [6]. Although a few physiotherapists talked about progressing the duration and intensity of aerobic exercise, they did not specifically mention progression to at least 20 minutes of aerobic exercise for their cases.

#### Outcome expectation of exercise

When physiotherapists were generally asked about their opinion on aerobic exercise after stroke, they expected aerobic exercise to benefit their patients. The outcomes they expected from aerobic exercise included improved general health, endurance, and motor function. Together with these expectations, some physiotherapists believed patients can be functionally too low level, or have severe cognitive or perceptual deficits to increase their heart rate to a sufficient intensity for cardiorespiratory benefit. Moreover, most physiotherapists reported that aerobic exercise is viewed as an ‘add-on’ or afterthought by physiotherapists at their centre. When this specific view was presented, often functional activities (e.g., walking, stair climbing) without a predefined intensity (see the next subtheme) were used with the hope to achieve cardiorespiratory benefits or to gradually improve endurance for their patients. Some believed that, because stroke patients’ cardiorespiratory fitness is typically very low, performing these short-duration and/or low-intensity functional activities will still improve their cardiorespiratory fitness and can be considered as performing aerobic exercise. Courtney, working in an inpatient unit, explains,

> I think when they’re that low level it, it does make a difference. 5 to 10 minutes at the end of a session when they’ve already done transfer training, gait training, pushing them that extra 5 to 10 minutes at the end, I think does make a difference when they’re that impaired going into it.

Conversely, other physiotherapists did not expect that the limited duration/intensity of aerobic exercise that they implemented during rehabilitation would provide cardiorespiratory benefits.

#### Monitoring and objective measures are not frequently used

In general, regardless of behaviour in relation to the use of aerobic exercise, few physiotherapists reported using objective measures to prescribe the intensity of exercise. Some physiotherapists used rate of perceived exertion [25] or general patient response to exercise (e.g., observing their breathing and facial expressions, ability to talk) to determine if the exercise was at an appropriate intensity. However, others did not mention intensity at all.

Limited use of objective measures to assess cardiorespiratory fitness and monitor exercise intensity likely resulted in patients being ‘under exercised,’ as noted by Alta, working in an outpatient unit,

> I probably am under exercising them, I’m guessing, like if I did the formal testing [submaximal exercise testing] I probably might push a little harder, but just in the time constraints we’ve got I feel like there’s so much other stuff to assess I really want to get them going fairly soon in their program and I feel like this has been a pretty sort of conservative so safe way to get them going on that exercise quickly and then build them up.

One physiotherapist thought formal exercise testing could be done more often; this physiotherapist tried an informal endurance test with one of their patients discussed. One physiotherapist mentioned having done sub-maximal exercise testing in the past but not anymore; in the case discussions, they mentioned doing a 6-minute walk test with one patient. Only one physiotherapist mentioned regularly doing 6-minute walking test or sub maximal exercise testing, and through the patient cases discussed how these exercise tests inform prescription (e.g., target heart rate). Lack of time was one of the reasons that many physiotherapists did not perform an objective assessment to determine the intensity of the aerobic exercise that patients should begin with.

*2) Profiles of people with stroke, as well as their goals and their exercise modality preferences Patients’ specific profiles influence aerobic exercise use*

Physiotherapists described various patient-related factors that facilitated or impeded use of aerobic exercise, including: patient education/understanding of the benefits of exercise, patient enjoyment of exercise, fatigue, patient functional level (often because the patient was considered too low level or too weak to be appropriate for aerobic exercise), already being active/fit (so aerobic exercise prescription was considered unnecessary), low/normal endurance, their pre-existing conditions and medical comorbidities (often cardiac), pain, fall risk, motivation, depression/anxiety, and their age. Some physiotherapists specifically mentioned patient’s (and their family’s) ‘buy in’ to the importance of exercise as being critical. Physiotherapists mentioned that patients generally complied with exercise, but a few refused; three of the patient cases discussed how exercise was refused due to fatigue, not seeing the benefit, and the belief that it would worsen their pain. Many physiotherapists indicated that some level of physical and cognitive function was necessary to be able to exercise safely (e.g., physically able to get on exercise equipment, low fall risk when walking, not ‘impulsive’). Therefore, if patients were severely affected cognitively and/or physically, they were usually not prescribed aerobic exercise. One physiotherapist discussed how patients’ functional levels can be insufficient to achieve heart rates at sufficient intensity for cardiorespiratory benefit.

Helen, working in an outpatient unit, elaborated,

> So barriers that we’ve encountered or that I’ve encountered that don’t enable us to achieve an exercise intensity that benefits the endurance is if they’re too weak, if they only have reliable activation of one side of their body, if they cannot physically move at a pace that’s fast enough to get their heart rate going high enough then it’s still better than not having it but it might not get us the results as effectively as someone who has full control of all four extremities.

Physiotherapists discussed patient safety while exercising in terms of risk of falls and cardiovascular events. Safety concerns often led to aerobic exercise without formal prescription or not being prescribed at all. A few physiotherapists discussed taking a gradual or cautious approach to exercise prescription (e.g., gradually ramping up intensity or duration of exercise) to ensure the patient is safe.

Most physiotherapists believed low tolerance early in rehabilitation leads to prescription of shorter duration exercise. However, this was mostly mentioned in response to general questions rather than in relation to specific patient cases. Generally, physiotherapists described prescribing exercise for ’higher functioning’ patients. Some physiotherapists prescribed aerobic exercise for patients who were sedentary or deconditioned, though it was not necessarily a patient-identified goal to improve endurance. Two physiotherapists mentioned that they prescribed exercise for patients who could exercise independently or could be supervised from a distance, so the patients could exercise outside of “rehabilitation intensity” time (see below), and they could make the most of limited time in therapy with the patients.

Pre-existing conditions, such as knee osteoarthritis, and some stroke complications (e.g., higher tone) and cardiovascular surgery were among the patient-related factors that led to modifying aerobic exercise prescriptions, and often prescribing aerobic exercise without specific intensity and or insufficient duration. Yvan mentioned,

> They were doing the MOTOmed consistently during the treatment sessions level one six minutes the first session that’s all they could tolerate. They couldn’t tolerate longer than six minutes. We started at a low duration, and they progressed, up to, level two ten minutes. [Level 1 and 2] are the intensity, so… it’s a very low level, like, zero is the motor takes over [and] does all the work whereas level one is the next one up from the motor taking over. With this patient, because they had that weakness in their leg it was mostly the motor doing the work for them, if it wasn’t their stronger leg pushing.
>
> Interviewer: so it was not really aerobic for this patient, right?
>
> Yvan: correct! it was aerobic equipment, yes,

Concerning the effect of patient age and exercise history, Courtney, working in an inpatient unit, said that “there is definitely a consideration of what their activity levels are prior because if it’s you know like a 90-year-old man who wasn’t very active before, our priorities for their exercise might be very different…. the age is something we consider and the type of activity levels they had prior [to their stroke].”

#### Patients’ goals, goal priorities, and preferences

Patient goals were mentioned frequently when discussing patient cases. Aerobic exercise tended to be used when patients had goals around improving endurance, walking distance/tolerance, reducing fatigue, and/or improving fatigue. Some patients expressed goals in such a way to imply improving aerobic capacity (e.g., to walk a flight of stairs without getting out of breath), and some patients had goals to return to their previous physical or sporting activities (e.g., walking program, playing basketball). When patients had the goal of improving endurance whether explicitly or implicitly, or they mentioned they were fatigued, aerobic exercise was usually not used during rehabilitation. Helen, working in an outpatient unit, described,

> So, this patient was prescribed the aerobic exercise. They were able to exercise at a reasonable intensity to get cardiorespiratory benefit. So, they identified a lot of goals that required just more physical endurance so for example, to be able to be move independent(ly), to be able to walk to the drug store, to be able to manage stairs, and things like that. It motivated me to ensure that the cardiorespiratory element was incorporated into the program.

Conversely, there was at least one case where a patient had a goal to improve endurance, but the physiotherapist did not employ aerobic exercise specifically because they perceived that it was not relevant to the patient’s rehabilitation goals. For example, rehabilitation goals for a patient were to increase strength in lower extremity, maximize independence for ambulation, transfers and stairs, improve balance and increase endurance but the physiotherapist did not prescribe aerobic exercise for them. This physiotherapist elaborated that “I felt like working on their balance to prevent falls as well as improving the way they were walking and transferring were more important to me so that they would be able to be safe when they went home”.

Most patients had goals related to improving mobility (e.g., independence, walking without an aid, stairs) rather than endurance. Particularly for lower functioning clients, patient rehabilitation goals were often to improve overall function (e.g., transfers, ambulation), rather than improve aerobic capacity, and physiotherapists focused on getting patients home safely. Goal priority was usually discussed concurrently with the barrier of time, in general questions and frequently for specific patients, as a barrier to aerobic exercise. Goal prioritization (other than safety) led to aerobic exercise being considered ’secondary’ to other goals, and therefore often being conducted without a specific prescription, or omitted altogether, due to the limited time that physiotherapists have with their patients.

Aerobic exercise use during rehabilitation was often influenced by what the patient wanted to do at home/after discharge. For example, one patient wanted to be able to return to work in which they had to be on their feet all day, and to lift at least ten pounds, because that was one of their work duties. Therefore, they practiced floor to waist lifting, waist to waist kind of carrying, rotations from one surface to another, lifting and carrying, walking around with ten-pound weights, for two hundred metres. Also, they went on the elliptical machine for 10 minutes, and then “NuStep” for 10-15 minutes, which means longer duration of aerobic exercise compared to the unit norms of maximum 10-15 minutes. Physiotherapists also reported that patient preference for different types of exercise equipment sometimes influences the equipment used in treatment. For example, Adele mentioned that two of her patients did not want to switch from the seated recumbent stepper to other equipment, like a treadmill, and this is why she continued to use the recumbent stepper with these patients.

#### Discharge setting of patients and their associated resources

Physiotherapists discussed the importance of patients continuing to exercise after they left rehabilitation. Most physiotherapists stated that discharge plan/destination often influences whether treatment plans during rehabilitation included aerobic exercise, and its specific prescription. Physiotherapists perceived that most patients have limited access to resources/equipment for ongoing exercise post-discharge (e.g., home exercise equipment). Some physiotherapists were mindful of this lack of resources/facilities, and prescribed exercises during rehabilitation that would be sustainable post-discharge (e.g., walking) but without specific intensity and or duration. As Syed, working in an inpatient unit, put it,

> I feel as though it’s probably more beneficial to have someone walk a half an hour every day as opposed to like, you know, maybe get motivated to do a forty-minute bike ride, at a higher intensity once every three weeks because it’s like too hard so I really try and do a graded approach because I think graded approaches are very, very effective, in terms of habit-forming behaviour especially when they’re going to go home and I’m not going to be there to be like “get on the bike, do it now”

Conversely, some physiotherapists noted that, because of limited resources/facilities in the community or at home (e.g., no program to refer patients to during COVID-19 and patients being discharged home where they do not have any aerobic equipment), the only aerobic exercise that patients would receive was during formal rehabilitation. Two inpatient physiotherapists mentioned that when it was not possible to continue doing aerobic exercise after discharge from their units, they would not prescribe it during rehabilitation either.

When patients had exercise equipment at home, some physiotherapists discussed how they would use similar equipment during rehabilitation to facilitate longer-term aerobic exercise participation post-discharge. On the other hand, one outpatient physiotherapist explained that, because their patients had treadmill at home and were already doing aerobic exercise, they did not duplicate this during rehabilitation and focused on other goals.

> I guess the only other people who I might not have that [do aerobic exercise] as part of their program, like during their session with me is people who are doing that stuff on their own anyway. Like some people have a treadmill at home and they’re already doing it and so I don’t necessarily duplicate that while they’re with us like I’ll work on something else instead because they’ve said that they’re already doing that.

In-patient physiotherapists often mentioned how, due to limited time to do aerobic exercise during rehabilitation, they communicated with out-patient therapists regarding following-up with aerobic exercise work that they had begun. In discussing individual cases, some physiotherapists mentioned referring their patients to cardiac rehabilitation.

*3) Influence of the health system priorities, rehabilitation intensity policy, and resources Access to resources*

physiotherapists reported that the availability of resources facilitated aerobic exercise prescription but when not available, they presented barriers. However, with few exceptions, resources were mostly discussed in the context of general barriers/facilitators, and not for specific patients. Most physiotherapists noted that they had plenty of exercise equipment available to them in their rehabilitation setting. However, physiotherapists also said that the equipment is a finite resource and needed to be shared with other physiotherapists. Having to share the equipment with other physiotherapists, and therefore only having access to the equipment for 10-15 minutes per patient session, was described as a reason for using aerobic exercise without specific intensity and/or duration when discussing specific patient cases. It was also mentioned as a barrier to prescribing aerobic exercise when physiotherapists were asked general questions. Adele, working in an inpatient unit, in response to “could you do maybe twenty to thirty minutes like, of aerobic exercise that exercise guidelines suggest?” explained,

> I could, but again, we were trying to balance out also sharing the equipment with other therapists, so I, that’s another restriction with resource.

And in response to general questions Yvan said,

> you know our gym is very busy, so it was hard to book the bicycle (for exercise testing) because you’re sharing with another program.

Therefore, collaboration between physiotherapists is required to ensure efficient and equitable use of limited resources.

Physiotherapists discussed how stroke patients often cannot use exercise equipment (e.g., due to motor impairment) and may not be safe or be able to walk fast enough to use walking for exercise, so it is useful to have adaptive equipment available for these patients. Therefore, most physiotherapists used the NuStep® (+/-leg stabilizers), though some also discussed arm ergometers. Two physiotherapists mentioned that patients can have joint pain with some exercise equipment, so it is useful to have a variety of options available to try. Physiotherapists discussed using treadmills or other devices with higher functioning patients, or transitioning to using a treadmill as patients’ function improves.

#### Policies and priorities

Ironically, the ‘rehabilitation intensity’ policy that is put in place to increase the amount of rehabilitation people with stroke receive [35], seemed to pose a significant barrier to the use of aerobic exercise. The Ontario Stroke Network has identified ‘rehabilitation intensity’ as a key indicator for evaluating system efficiency and effectiveness [35]. Rehabilitation intensity entails an individualized treatment plan that involves a minimum 3 hours of direct task-specific therapy per patient per day by the core therapies (i.e., physiotherapy, occupational therapy, and speech-language pathology) for at least six days per week. The task-specific therapies that are ‘counted’ towards rehabilitation intensity must be individualized (i.e., group therapy is not included) and a maximum of 33% of therapy time can be with therapy assistants [35]. It is important to note that ‘intensity’ of rehabilitation, within the context of the Rehabilitation Intensity policy, refers only to the amount of time patients spend in therapy. This contrasts with ‘intensity’ of exercise prescription, which is defined as the rate of metabolic energy demand during exercise that relates to the level of challenge or difficulty of the exercise [36]. Some physiotherapists were of the impression that aerobic exercise using equipment (e.g., NuStep, treadmill) does not count towards rehabilitation intensity, whereas others thought it did count, provided the exercise was conducted 1:1 with the physiotherapist. What counted and did not count as rehabilitation intensity seemed to differ by institution. Rehabilitation intensity was always raised in the context of general barriers and facilitators, but not when discussing specific cases. Two physiotherapists talked about aerobic exercise not being incentivized by current health system policies. Jennifer, working in an inpatient unit, explained,

> so I guess the biggest thing is that it [aerobic exercise] is not incentivized here with our metrics that I mentioned before with our rehab intensity.

Physiotherapists who talked about rehabilitation intensity as a barrier in general questions, when discussing specific cases usually employed aerobic exercise without specific exercise intensity and or duration. Additionally, some asked the patient to come to the gym outside of their formal therapy time, as a strategy to implement some aerobic exercise outside the constraints of this healthcare policy in Ontario. One physiotherapist reported that they ignore the management’s enforcement of this healthcare policy because they deem using aerobic exercise equipment for people with stroke necessary. Alternatively, this lack of behavioural control led some physiotherapists to not use aerobic exercise at all due to the rehabilitation intensity policy.

Relatively short lengths of stay, which is also dictated by health system policy, limits how much aerobic exercise a physiotherapist can incorporate into their treatment. However, one physiotherapist indicated that they expect aerobic exercise to support improving function, even if they are not working on activities like transfers and ambulation directly.

Courtney, working part-time in an inpatient unit, said,

> I think a lot of it is just this, focus on function, we really kind of live or die by the FIM scores … to show the most recovery for anybody, it’s maximizing independence on transfers and ambulation, and though we know that aerobic exercise is important for helping those things, I think …it feels like if we’re not working on transfers and ambulation, we’re not improving it even though improving cardiovascular fitness helps with [function].

One physiotherapist noted changes in stroke rehabilitation care; that is, patients were increasingly being admitted to rehabilitation earlier post-stroke than previously. This translated to patients not being ready to be tested for and being prescribed aerobic exercise (e.g., too early after cardiac surgery, too weak to pedal fast enough to get any cardiorespiratory effect).

#### Challenges and opportunities of providing care in a multidisciplinary team

Physiotherapists discussed providing care in the context of a multi-disciplinary team. While aerobic exercise prescription falls within the scope of practice of physiotherapists [8], often having the support/buy-in from other team members (e.g., interprofessional collaboration with the management, physicians, nurses) was important. This can present a barrier if these team members are not supportive, but a facilitator when they are. In response to the general question of what are the factors that helped you to implement aerobic exercise during stroke rehabilitation, Marianne explained,

> Having access to a physician who’s well-versed in yes’s and no’s like, and they’ll, they want to have the conversation so, I can easily go to the hospitalist on the unit and say, “do you think it would be okay for me to proceed with this?” our physiatrists are really supportive.

Physiotherapists also discussed intraprofessional collaboration in the context of continuity of care/discharge planning, e.g., an in-patient physiotherapist can do exercise testing and refer to out-patient physiotherapist for implementing the prescription, or to cardiac rehabilitation.

As Marianne, working in an inpatient unit, explained,

> Some people leave here with a referral, recommendation that they should be referred to cardiac rehab. So, we rely on our colleagues in cardiac rehab now because they’ll accept the people with stroke.

#### Group norms regarding aerobic exercise assessment and implementation

Some physiotherapists discussed group norms on their units in response to general questions asked during the interview. These norms included: other physiotherapists not doing exercise testing, sharing equipment with other patients and therapists (i.e., therefore not using one equipment for more than 10-15 minutes), gait and transfer training being the main focus of physiotherapy on the unit, doing/not doing aerobic exercise, aerobic exercise being an add-on toward the end of the session, and not monitoring heart rate. These norms usually led to implementing aerobic exercise without specific intensity and/or duration. Alta, working in an outpatient unit, explained,

> “I haven’t seen anybody else do it [heart rate monitoring] in our clinic. So it was, it’s not sort of always on my radar as necessary.”

Keeping the status quo of the unit or using the same method that the physiotherapist has always used appeared to be a common barrier for trying new methods or applying best practice guidelines (e.g., performing submaximal exercise testing, prescribing aerobic exercise with specific intensity and duration).

### Influence of COVID-19 pandemic

As the data collection happened during COVID-19 pandemic, it greatly exacerbated the challenges that already existed for prescribing aerobic exercise to patients post stroke. For example, equipment needed to be spaced out in the gym as per infection control policies, and this translated into having less equipment available as the space was large not enough to hold in all the devices with the required distance. Additionally, managing COVID-19 cases required hospitals to redeploy many healthcare providers to other units. This meant that the number of physiotherapists available in their previous positions decreased, and the caseloads of those staying in the stroke rehabilitation units increased. This in turn led to even further limitations in time during therapy sessions.

The priorities of the healthcare system also shifted toward earlier discharge of non-COVID patients, which changed the goal priorities and discharge plans of the stroke rehabilitation units. These units mostly focused on sending patients home as soon as possible. Yvan, working in an inpatient unit, stated,

> you have different role than you would before COVID hit, on some days, some days you know you’re a stroke physio and you’re seeing your patients as you normally would but there’s days where you know the priority is not stroke rehab so you kind of have to adapt and then what you do for the patients is what you have to absolutely do for the patients and I would say aerobic training really falls by the wayside because we’re looking at how do you get the patient home safely, quickly.

Continuity of care after inpatient stroke rehabilitation was mostly lost. Yvan commented,

> and then there’s nothing to refer to them, you know with COVID out-patients is virtual, you know, all the YMCA programs, that you know [specific community program] you know shut down, you know just those programs that were you know like mall walking that sort of thing, all that stuff was shut down so, you know the patients really have no options.

Treatment plans were also affected. Syed, working in an inpatient unit, mentioned,

> Not only having a stroke but, you know, hypertension and all these other things that puts you at a higher, like COVID risk, many people didn’t have the resources to, you know, go on a bike after they got out of here, so I started them on graded (functional activities like walking) because I wanted them to be able to continue the exercise in a graded manner with what they were able to do, and or even financially afford out, when they were in the community.

Patients’ goals were sometimes affected because their families were overprotective (i.e., limiting their activities in the community during the pandemic) during the pandemic. For example, as Adele, working in an outpatient unit, stated,

> again, some of these patients used to have goals of, you know, being able to grocery shop and then now they, the family will refuse to let them go out, ‘cause I find grocery shopping, you know, being in a grocery store where you have to walk further, pushing a cart, again, that’s another kind of test for endurance, but again, family has taken that away from some patients.

## DISCUSSION

In this study, we aimed to obtain a better understanding of physiotherapists’ use of aerobic exercise during stroke rehabilitation for each specific case with stroke. We identified 3 main themes for this study, namely, physiotherapists’ perspectives and practices around aerobic exercise; profiles of people with stroke, as well as their goals and their exercise modality preferences; and influence of the health system priorities, rehabilitation intensity policy, and resources.

To our knowledge, this is the first study exploring physiotherapists’ clinical reasoning for aerobic exercise use **during stroke rehabilitation** by discussing the **specific patient cases**. In this study, we were able to capture the perspectives of both in-patient and out-patient physiotherapists in our interviews and provide a more comprehensive view of the perspectives of physiotherapists working at stroke rehabilitation. Our participants showed varying behaviours in relation to aerobic exercise use: not employing it at all, putting people with stroke on ‘aerobic’ equipment as warm up or cool down only, using low-intensity functional activities expecting that this will improve cardiorespiratory fitness, and prescribing and implementing aerobic exercise with specific duration and intensity (with or without prior objective testing). Based on these findings, the behaviour regarding the use of aerobic exercise is better understood on a continuum rather than being a binary behaviour of prescribing or not prescribing aerobic exercise. As mentioned before, 77% of physiotherapists working in an adult neurological rehabilitation settings across Canada stated they prescribe aerobic exercise to their clients with stroke [8]. However, combining this with the aerobic exercise usage behaviour observed in our study, some of the physiotherapists who filled this survey may have counted aerobic activities without specific intensity, frequency and/or duration as formal aerobic exercise prescription. Similar to our study, Doyle and MacKay-Lyons report that unofficial methods such as observing patient responses to exercise, the patient feedback, and/or using rating of perceived exertion and/or the Talk Test are the most frequently used methods for determining the intensity of aerobic exercise [8]. Only 30% or fewer of the physiotherapists indicated they calculated the heart rate target (standard practice[6]) of the patients for performing aerobic exercise [8]. Therefore, the proportion of physiotherapists who are prescribing optimal aerobic exercise during stroke rehabilitation may be lower than 77%.

This study revealed that barriers mentioned in response to general questions asked from physiotherapists differ from the ones raised when discussing specific patient cases. Profiles of the people with stroke, as well as their goals were repeatedly discussed by the physiotherapist as an important consideration when using aerobic exercise in daily practice. A similar recent study showed that health care professionals take goals of people with stroke into consideration when planning their treatments; which usually leads to aerobic exercise having lower priority than other interventions [15]. This is because people with stroke prioritize improvement in function especially during inpatient rehabilitation to other goals [15]. Patient-related factors (e.g., cardiac risks, cognitive/perceptual deficits, balance impairments, lack of motivation and fatigue) are commonly reported as barriers to aerobic exercise use post-stroke in previous studies [8,37,38]. However, the importance of patients’ preferences and exercise history that were found in our study have rarely been reported in previous literature[39,40]. We found that what the patient wanted to do at home/after discharge also influences prescription of aerobic exercise during rehabilitation. Tailoring exercise programs to each individual improves the efficacy of interventions for increasing physical activity in people with stroke [41], and catering to patient preferences may then help to increase longer-term uptake of physical activity and exercise. The emergence of patient’s preference in our study may have been facilitated by the use of chart-stimulated recall, which tends to highlight the less objective aspects of care such as assessment of patients’ life expectancy [16].

In this study, patient motivation, education/understanding of the benefits of exercise, enjoyment of exercise, perceptions of the benefits of exercise, exercise history, and age were introduced as factors facilitating usage of aerobic exercise during rehabilitation. Conversely, being already active/fit was one of the factors that led to no aerobic exercise prescription during rehabilitation post stroke. This means even if physiotherapists are not concerned for the safety of their patients (related to the patient’s cardiac, physical or cognitive impairment) they still may not use aerobic exercise as they deem it to be unnecessary for these patients. However, previous research shows that aerobic training has a significant positive effect on risk factors for recurrent stroke (i.e., systolic blood pressure and fasting glucose) [42]. Therefore, using aerobic exercise, preferably with supervision, for already active people with stroke could help reduce the long-term stroke risks for these patients. People with minor stroke have a preference to exercise under the supervision of a healthcare professional in order to keep being physically active following discharge [43]. A study aiming to compare the effects of supervised versus unsupervised exercise programs for ambulatory stroke survivors reported that, although both supervised and unsupervised exercise programs for people with stroke can lead to increased walking speed (retained for 1 year), the supervised program was superior [44].

The theme of policies put in place by the healthcare system (e.g., rehabilitation intensity) was the most commonly discussed barrier that was only mentioned in response to **general questions**. The rehabilitation intensity policy was originally implemented because previous research showed that adequate duration of therapy during rehabilitation is associated with successful inpatient rehabilitation outcomes (e.g., increase in FIM and activity in people with stroke) [45–47]. Roberts et al., recently showed that increased physiotherapy time in rehabilitation resulted in significantly higher FIM gains post-stroke [48]; an additional hour of physiotherapy per day was associated with an increase in FIM scores of 7.6 in this inpatient rehabilitation program, which mostly consisted of people with a neurological diagnosis [48]. Rehabilitation intensity of 3 hours per day of physiotherapy, occupational therapy and speech therapy is also used in Japanese stroke rehabilitation in Kaifukuki (convalescent) rehabilitation wards [49].

In our study, two physiotherapists specifically mentioned that aerobic exercise is not incentivized in the current model of care with the use of rehabilitation intensity, and if it was, aerobic exercise would be done more often and better (i.e., with more formal prescription). Additionally, we found that there was no consensus among physiotherapists from different sites as to what counts toward rehabilitation intensity. Lack of supportive policies in the use of clinical practice guidelines has also been shown to be a major barrier to implementation of guidelines for the management of people with hypertension as it affects the healthcare providers’ awareness, adherence and attitudes to the use of guidelines [50].

Following best practice guidelines (e.g., for aerobic exercise during rehabilitation post stroke) becomes more challenging when the mandated policies and funding models do not support them. For stroke, hospital funding is partially dependent upon changes in Functional Independence Measure (FIM[51]) scores. This focus on needing to improve FIM scores leads to exercise that targets functional activities rather than intense aerobic exercise. Another recent study also found that when reimbursement policies are not consistent with the clinical practice guidelines for breast cancer treatment in Europe, it will lead to the inability to clinically implement the best practice guidelines for the patient with breast cancer [52]. This poor alignment acts as a barrier and disincentive to the best breast cancer care in these countries [52].

The other barrier mostly discussed in response to **general questions** was limited resources and having to share equipment with other physiotherapists. Both in-patient and out-patient physiotherapists talked about their patients being functionally low level, and therefore, believed that a seated recumbent stepper is the safest option for these patients. The combination of limited resources and low patient level meant that they had a norm in their units to not put their patients on the recumbent stepper for more than a short amount of time. Limited resources is frequently cited as one of the factors negatively influencing use of aerobic exercise post stroke [8,10,14,15]. Goal priority, simultaneously with the barrier of time limitation, were discussed in response to both the general and the patient specific questions. Research findings from a scoping review on the barriers and facilitators to applying patient-centred goal-setting practice in rehabilitation suggest that one of the most common barriers to person-centred goal setting in rehabilitation is the insufficient time for providers [53]. Although all the physiotherapists had set rehabilitation goals with their patients, lack of time led to postponing some goals to a later time during their rehabilitation and instead focusing on the ones that the physiotherapist deemed more important for the patients’ safe and quicker discharge. Some in-patient physiotherapists had only 45 minutes per session with some of their clients and believed that if they had one hour, they could do aerobic exercise. However, most out-patient physiotherapists who had one hour with their patients still could not do more than 20 minutes of aerobic exercise due to lack of equipment and sharing equipment with other therapists. In a country-wide survey study conducted in the Netherlands, the minimum time dedicated to exercise therapy per weekday for people with stroke was reported to be 22 minutes while the hospital policy recommended 24 minutes, and the Dutch Clinical Practice Guidelines for Patients with Stroke recommended this amount to be 40 minutes per day [54]. In this previous study, the physiotherapists perceived that they had insufficient time within therapy to comply with the clinical guidelines [54].

Keeping the status quo of the unit or continuing to use the same method for different patients acted as barriers to performing an objective testing or to prescribing a formal aerobic exercise for each case. This is called passivity in the provision of care, which is usually better seen in qualitative studies that use chart stimulated recall, rather than conventional qualitative interviews where general questions are asked [16]. Institutional/unit culture can act as a barrier to early mobilization of patients in an intensive care unit [55]; specifically the presence of an endotracheal tube was perceived as a barrier to mobilization by many staff, although it was not a contraindication [55]. A systematic review of the factors influencing physical activity and rehabilitation among survivors of critical illness found that one of the major themes is culture and team influences, including leadership, interprofessional communication, administrative buy-in, clinician expertise and knowledge [56]. Challenges and opportunities of intra-and inter-professional collaboration was observed in our study too. Our study showed that because physiotherapists provide care in the context of a multi-disciplinary team, having the support/buy-in from other team members (e.g., management, physicians, nurses) can facilitate aerobic exercise prescription. Continuing education and advocacy about the benefits and safety of aerobic exercise for people with stroke may be required to get this ‘buy-in’ from non-physiotherapist healthcare providers.

Limited use of objective measures to assess cardiorespiratory fitness and monitor the exercise intensity likely led to the use of functional exercises instead of aerobic exercise. Additionally, physiotherapists believed that these exercises could still improve cardiorespiratory fitness of people with stroke. However, a study by MacKay-Lyons and Makrides showed that when aerobic conditioning is not a major component of the stroke rehabilitation program, the patient’s heart rate is below 40% of the heart rate reserve for 95% of the total time spent in physiotherapy between 2-14 weeks post-stroke [57]. A more recent study confirmed these findings by showing that only 4 of 19 participants in inpatient stroke rehabilitation reached 30–45% of heart rate reserve during physiotherapy sessions [58]. In our study, there was a perception that having a group exercise program (which was active in all the settings), in addition to 1:1 physiotherapy sessions, would remove many of the barriers related to physiotherapists’ clinical considerations, health system policies and priorities, and organizational resources. The group exercise program comprised of a supervised program delivered in a group setting that was available to patients 3 times per week [9]. The primary treating physiotherapist conducted an exercise test and referred their patients to the group with an exercise prescription and progression guidelines [9]. Availability of the group allowed treating physiotherapists to focus their 1:1 time with their patients on priorities other than cardiorespiratory fitness. While these groups were paused early after the COVID-19 pandemic, they were technically available to physiotherapists at the time of the interviews. Even prior to the pandemic, referral to the fitness groups was low (<10% of patients). Therefore, while several physiotherapists in our study specifically stated that such a group would address many of their barriers to use of aerobic exercise (e.g., limited time and resources), low uptake of the fitness groups contradicts this view. Our experiences with implementing the fitness groups [9,23] combined with the results of this study, suggest that patient-related factors (e.g., co-morbidities) and lack of supportive policies for aerobic exercise use represent significant challenges to the use of aerobic exercise during stroke rehabilitation.

### Limitations

As mentioned before, the four stroke rehabilitation programs that we recruited from were participating in another research study [23], where physiotherapists were trained in aerobic capacity testing and exercise prescription for people with stroke. Consequently, these physiotherapists had expertise in using aerobic exercise for persons with stroke. The results may not be transferable to other stroke settings, which have not received the same training. However, conducting the study in this setting allowed us to identify factors preventing conformation to best practice guidelines where common barriers (e.g., limited time’), should have been addressed, at least in theory.

This study was conducted during the COVID-19 pandemic. Several physiotherapists who would have been eligible for this study were redeployed to other units in the hospital in response to the pandemic. Therefore, it was not possible for them to participate in the study. Similarly, some physiotherapists declined to participate due to feeling overwhelmed because of the pandemic. Consequently, the number of physiotherapists interviewed in this study was less than anticipated. However, as explained above, the information power of the study was still sufficient.

Although CSR is mainly used to diminish the effect of recall bias, clinicians may have not accurately recalled their decision making-processes, so they may have retrospectively provided an explanation for their clinical decision making [16]. To mitigate this issue, we asked the participants to bring the charts of their four recently discharged patients to shorten the duration between the encounter and the interviews and, therefore, facilitate their recall. Another bias that may play a role in this study is selection bias [16]. Physiotherapists may have had more than 4 recently discharged patients, and they may have inadvertently chosen the charts that had more details included. We tried to mitigate the effect of researcher’s biases in two ways: firstly, by having researchers with different backgrounds code and analyse the data separately; secondly, the interviewer kept notes of their encounters and interviews with the participants and the potential impact they may have had on the interviews.

## Conclusion

This study explored physiotherapists’ use of aerobic exercise for people with stroke and helped us to obtain a better understanding of their clinical reasoning and factors that affect their decisions for each specific case. We found that in day to day practice, specific profiles and goals of people with stroke are at the centre of physiotherapists’ reasoning for aerobic exercise prescription or lack thereof. The policy put in place by the healthcare system in Ontario (i.e., rehabilitation intensity) may not be supportive of the implementation of best clinical practice guidelines for prescribing aerobic exercise to people with stroke during rehabilitation. Organizations can facilitate aerobic exercise prescription by improving their culture of working among multidisciplinary teams and reevaluating the existing group norms that prevent physiotherapists from adherence to the practice guidelines.

## Data Availability

Raw data are not available.

## Acknowledgements

The authors would like to thank Devanshi Joshi, Saira Thavaneethan, and Sarah Thompson for their help during the interviews and for transcribing the interviews.

## APPENDIX 1 Interview guide for physiotherapists

*Interviewer will first remind participants about the purpose of the study, confidentiality, and consent to be audio recorded before proceeding with the questions below*.

*COM-B items probed with individual questions: C=Capability, M=Motivation, O=Opportunity, B=Behaviour*

I am going to ask you questions about your experiences with implementing aerobic exercise during stroke rehabilitation. You are free to skip any questions that you do not want to answer, and you can end the interview at any time.

We will start the interview with some general questions, and then we will discuss the patient charts that I asked you to prepare.

For the purpose of this discussion, we define aerobic exercise as planned, structured, and repetitive physical activity performed with a prescribed frequency, duration, and intensity aimed to improve or maintain cardiorespiratory fitness.

Are you ready to begin?

1. Tell me about your understanding of aerobic exercise after stroke. [**C**]
2. What have your experiences been with implementing aerobic exercise for your clients with stroke? [**B**]

*Prompts:*

Does your setting incorporate aerobic exercise? [**O**]

Do you incorporate into your practice? If so, when did you start? Why did you start? [**B**] Who is involved in supporting this practice? [**O**,**B**]

How is it implemented? [**O**,**B**]

How does aerobic exercise benefit patients (if at all) early after stroke? [**M**]

3. What are some of the challenges you’ve experienced with implementing aerobic exercise during stroke rehabilitation?
4. What things have helped you to implement aerobic exercise in stroke rehabilitation?

*Prompts*

What kind of education or training have you received related to aerobic exercise post-stroke? How did this training adequately prepare you (or not) for the types of patients you encounter in your practice? [**C**]

Do you routinely determine if each of your patients is suitable for aerobic? How do you decide whether to prescribe aerobic exercise for individual patients? [**M**,**B**]

Is there anything within your practice setting that facilitates or prevents you from completing aerobic exercise with your patients? [**O**]

Are there any other external factors (i.e., outside of your own knowledge, skills, or beliefs, or your practice setting) that influence your ability to conduct aerobic exercise with your clients? [**O**]

*Chart-stimulated recall*

5. Tell me a little bit about this patient, without providing any identifying information (e.g., their history, goals in rehabilitation, functional limitations or key impairments that you addressed in your treatment plan, and the patient’s progress). [Patient **C**]
6. Was this patient prescribed aerobic exercise as part of their rehabilitation? [Physiotherapist **B**, Patient **O**]
7. *If they were prescribed exercise*: what were some of the factors that influenced your decision to incorporate aerobic exercise into their rehabilitation program?
8. *If they were not prescribed exercise*: what were some of the factors that influenced your decision to not incorporate aerobic exercise into this patient’s rehabilitation program.

*Prompts*

*Ask about patient-specific factors (risks with exercise, comorbidity, medications) [Patient **C**] (patients’ personal characteristics, patient goals) [Patient **M**], clinician-specific (knowledge, skills) [Physiotherapist **C**], perceived organizational barriers (patient length of stay, organizational priorities, support required of other healthcare professionals or leaders, access to adapted exercise equipment) [Physiotherapist **O**], barriers of innovation (e.g. gaps in research knowledge specific to complex patients) [Physiotherapist **C**]*

9. Would you describe this patient case as ‘typical’ or ‘unique’? What characteristics are common between this patient and other patients that you treat? What characteristics are less common/unique?
10. Do you use a similar approach to prescribing aerobic exercise do other patients as you did with this patient? Why?

*Repeat questions 4-8, as appropriate, for all charts*.

11. Having now reviewed all the charts, what are the similarities and differences in your approaches to aerobic exercise prescription between these patients? What are the reasons for these differences/similarities?

*Questions modified from: Pak et al., 2015; AB et al., 2009; Rochefort et al., 2012*

## Notes

### Competing Interest Statement

The authors have declared no competing interest.

### Author Declarations

The Research Ethics Board of the University Health Network gave ethical approval for this work (protocol number: 20-5695). The Research Ethics Board of Sunnybrook Research Institute gave ethical approval for this work (protocol number: 3605). The Research Ethics Board of Hamilton Health Sciences gave ethical approval for this work (protocol number: 11523)

